# Dissemination and co-circulation of SARS-CoV2 subclades exhibiting enhanced transmission associated with increased mortality in Western Europe and the United States

**DOI:** 10.1101/2020.07.13.20152959

**Authors:** Yuan Hu, Lee W. Riley

## Abstract

Mechanisms underlying the acute respiratory distress syndrome (ARDS)-like clinical manifestations leading to deaths in patients who develop COVID-19 remain uncharacterized. While multiple factors could influence these clinical outcomes, we explored if differences in transmissibility and pathogenicity of SARS-CoV2 variants could contribute to these terminal clinical consequences of COVID-19. We analyzed 34,412 SARS-CoV2 sequences deposited in the Global Initiative for Sharing All Influenza Data (GISAID) SARS-CoV2 sequence database to determine if regional differences in circulating strain variants correlated with increased mortality in Europe, the United States, and California. We found two subclades descending from the Wuhan HU-1 strain that rapidly became dominant in Western Europe and the United States. These variants contained nonsynonymous nucleotide mutations in the Orf1ab segment encoding RNA-dependent RNA polymerase (C14408T), the spike protein gene (A23403G), and Orf1a (G25563T), which resulted in non-conservative amino acid substitutions P323L, D614G, and Q57H, respectively. In Western Europe, the A23403G-C14408T subclade dominated, while in the US, the A23403G-C14408T-G25563T mutant became the dominant strain in New York and parts of California. The high cumulative frequencies of both subclades showed inconsistent but significant association with high cumulative CFRs in some of the regions. When the frequencies of the subclades were analyzed by their 7-day moving averages across each epidemic, we found co-circulation of both subclades to temporally correlate with peak mortality periods. We postulate that in areas with high numbers of these co-circulating subclades, a person may get serially infected. The second infection may trigger a hyperinflammatory response similar to the antibody-dependent enhancement (ADE) response, which could explain the ARDS-like manifestations observed in people with co-morbidity, who may not mount sufficient levels of neutralizing antibodies against the first infection. Further studies are necessary but the implication of such a mechanism will need to be considered for all current COVID-19 vaccine designs.

## Introduction

Severe acute respiratory syndrome coronavirus-2 (SARS-CoV2) is the third highly-pathogenic betacoronavirus to cause epidemics in multiple countries this century, after SARS-CoV1 in 2002-3 and the Middle East respiratory syndrome coronavirus (MERS-CoV) in 2012. The pandemic of COVID-19, the disease caused by SARS-CoV2, began in Wuhan, China in December 2019 (1), and by end of June 2020, more than 10 million cases and 500,000 deaths have been reported worldwide. Based on the reported number of cases and deaths at the end of the first week of June 2020, the cumulative case-fatality rates (CFR) in Western European countries ranged from nearly 10% in Spain to 15% in France (https://www.worldometers.info/coronavirus/). However, Germany, despite reporting the eighth highest number of cases in the world, had a CFR of 4.7%, the lowest in Western Europe. In the United States, the first cases of COVID-19 were recognized in Washington state and California in January 2020 (2). The cumulative CFR in the United States at the beginning of June was 5.6%. New York City became the epicenter of the epidemic in North America in early April. By end of May, the cumulative CFR in the five boroughs of New York City reached 10%. These regional differences in mortality have been attributed to factors such as the timing of the implementation of social distancing measures, differences in population age structure, the number of tests performed per population, prevalence of co-morbidities, regional differences in clinical care (e.g., the number of intensive care units per population), as well as socioeconomic and environmental factors. One factor that has not yet received wide attention is the effect on mortality of circulating strain variants of SARS-CoV2.

The early reports of SARS-CoV2 genome sequence analyses have noted slow progressive accumulation of single nucleotide mutations relative to the reference strain from Wuhan (HU-1, MN908947)(3-6). Its low sequence diversity was thought not to overtly affect viral function or pathogenicity. However, a more recent study using ultra-deep sequencing (average coverage exceeding 2,000,000 X) has shown that nucleotide variants of the virus may be more common than previously recognized and that even mixed populations (quasispecies) of the virus could occur in a patient with COVID-19 (7). A study of only 11 viral isolates from 11 COVID-19 patients in Zhejiang, China identified over a 2-week period revealed 33 distinct mutations in the coding regions of the virus (7). These strain variants included several founding subclades of SARS-CoV2 now circulating in different regions of the world. They further observed that these variants exhibited as much as 270-fold differences in viral load and cytopathic effect (CPE) in Vero-E6 cells infected with the virus (7). Thus, it is beginning to be recognized that viral mutations can affect virus function, at least in vitro. In fact, a recent report described global dissemination of a SARS-CoV-2 variant with a mutation in the spike protein (D614G), suggesting that this strain gained fitness advantage for increased infectivity (3).

The observed geographic differences in COVID-19 CFR led us to speculate that, in addition to increased transmissibility, viral mutations may contribute to differences in mortality. We analyzed 34,412 SARS-CoV2 sequences deposited in the Global Initiative for Sharing All Influenza Data (GISAID) SARS-CoV2 sequence database (https://www.gisaid.org/) (as of June 4, 2020) to assess if regional differences in circulating strain variants correlate with differences in mortality in Europe, United States, and California. We found several sequences with missense mutations in Orf1ab, spike protein, and Orf3a genes in strains that were observed to dominate in Europe and the United States, associated with high CFR regions, controlled for the timing of initiation of social-distancing measures, population age structure, number of tests/population performed, prevalence of co-morbidities, and number of hospital beds/population. These observations have implications for predicting the course of the pandemic as well as in vaccine design and safety.

## Results

We compared CFRs by 1) country in Western Europe, 2) state in the US, and 3) county in California (Table 1). The cumulative CFRs were calculated based on COVID-19 cumulative number of deaths divided by the cumulative number of cases as reported by Worldometer’s database on June 6, 2020 (https://www.worldometers.info/coronavirus/). In Western Europe, we included countries that reported >150,000 cases (Italy, Spain, France, Germany, and UK), while in the US, we included states that reported >100,000 as of May 31 (New York, New Jersey, Illinois, California, Massachusetts). In California, we included counties from which > 90 SARS-CoV2 sequences were deposited in the GISAID database (San Francisco, Santa Clara, and San Diego county). In Western Europe, Germany had the lowest CFR at 4.7% compared to >10% in the other countries. In the US, California reported the lowest CFR at 3.4%, while New York had a CFR of 7.7%; the five boroughs of New York City reported a CFR of 10%. In California, Santa Clara county reported a CFR of 5.1%, while San Francisco reported a CFR of only 1.6% (Table 1).

**Table 1.** Cumulative case-fatality rates (CFR) due to COVID-19 as of June 6, 2020 in Western Europe, US, and California by dates of initiation of lockdowns, median age, co-morbidities, number of hospital beds/1000 population, and number of RT-PCR tests performed/1 million population. The countries in Western Europe included those that reported >150,000 cases, while the states included those that reported >100,000 cases. The California counties included those that submitted >90 SARS-Co2 sequences to the GISAID database.

| Region | CFR (%)<sup>1</sup> | Lockdown initiation date<sup>2</sup> | Median Age (yrs)<sup>3</sup> | Co-morbidities (DM%<sup>4</sup> Htn%<sup>4</sup>) |  | Hospital beds /1000 pop<sup>5</sup> | Number of PCR tests/1 M pop<sup>1</sup> |
| --- | --- | --- | --- | --- | --- | --- | --- |
| **Western Europe** |  |  |  |  |  |  |  |
| Germany | 4.7 | 2020/3/22 | 47.8 | 10.4 | 28 | 8 | 51,918 |
| Spain | 10 | 2020/3/14 | 43.9 | 6.9 | 25 | 3 | 86,921 |
| UK | 14 | 2020/3/23 | 40.6 | 3.9 | 20 | 2.5 | 73,763 |
| Italy | 14 | 2020/2/21 | 46.5 | 5.0 | 30 | 3.2 | 66,970 |
| France | 15 | 2020/3/17 | 41.7 | 4.8 | 29 | 6 | 21,216 |
| **United States** | 5.6 | -- | 38.2 | 10.8 | 32.1 | 2.8 | 60,944 |
| New York | 7.7 | 2020/3/22 | 38.7 | 9.4 | 29.4 | 2.7 | 121,289 |
| New Jersey | 6.9 | 2020/3/21 | 39.8 | 8.1 | 33 | 2.4 | 103,453 |
| Illinois | 4.4 | 2020/3/21 | 37.9 | 9.5 | 32.2 | 2.5 | 78,988 |
| California | 3.4 | 2020/3/19 | 36.1 | 9.7 | 28.5 | 1.8 | 56,652 |
| Massachusetts | 7.1 | 2020/3/24 | 39.4 | 8.3 | 28.6 | 2.3 | 98,740 |
| **California county** |  |  |  |  |  |  |  |
| San Francisco | 1.6 | 2020/3/17 | 38.2 | 6.7 | 29.6 | 6.4 | 94,870 |
| Santa Clara | 5.1 | 2020/3/19 | 33.9 | 7.3 | 30.1 | 5.2 | 46,555 |
| Los Angeles | 4.3 | 2020/3/19 | 37.5 | 8.1 | 30.3 | 7.1 | 68,860 |
| San Diego | 4 | 2020/3/19 | 35.6 | 6.9 | 29.5 | 5.5 | 57,400 |

<sup>1</sup>Worldometer, https://www.worldometers.info/coronavirus/
<sup>2</sup>https://www.nbcnews.com/health/health-news/here-are-stay-home-orders-across-country-n1168736
<sup>3</sup>https://www.cia.gov/library/publications/the-world-factbook/fields/343.html; https://worldpopulationreview.com/
<sup>4</sup>https://www.diabetesatlas.org/upload/resources/2019/IDF_Atlas_9th_Edition_2019.pdf (Prevalence of diabetes [DM], % of population ages 20 to 79 in 2019, age-adjusted); https://gis.cdc.gov/grasp/diabetes/DiabetesAtlas.html# (Prevalence of DM, adults >18 years, 2016, USA); https://www.who.int/nmh/countries/; https://gis.cdc.gov/grasp/diabetes/DiabetesAtlas.html# (Prevalence of hypertension [Htn] in adults >18 years in 2015-17, world);
[https://nccd.cdc.gov/DHDSP\_DTM/rdPage.aspx?rdReport=DHDSP\_DTM.ExploreByLocation&rdProcessAction=&SaveFileGenerated=1&isLocation=06&irbFilterby=1&isPriority=P3&isTopic=T9&hidLocation=06&hidTopic=T9&hidTopicName=Hypertension&isYear=9999&hidYear=9999&irbShowFootnotes=Show&rdICL-iclIndicators=BR011%2cBR012&iclIndicators\_rdExpandedCollapsedHistory=&iclIndicators=BR011%2cBR012&hidPreviouslySelectedIndicators=&DashboardColumnCount=1&rdShowElementHistory=divTopicUpdating%3dHide%2cisTopic%3dShow%2c&rdScrollX=0&rdScrollY=0&rdRnd=22817](https://nccd.cdc.gov/DHDSP_DTM/rdPage.aspx?rdReport=DHDSP_DTM.ExploreByLocation&rdProcessAction=&SaveFileGenerated=1&isLocation=06&irbFilterby=1&isPriority=P3&isTopic=T9&hidLocation=06&hidTopic=T9&hidTopicName=Hypertension&isYear=9999&hidYear=9999&irbShowFootnotes=Show&rdICL-iclIndicators=BR011%2cBR012&iclIndicators_rdExpandedCollapsedHistory=&iclIndicators=BR011%2cBR012&hidPreviouslySelectedIndicators=&DashboardColumnCount=1&rdShowElementHistory=divTopicUpdating%3dHide%2cisTopic%3dShow%2c&rdScrollX=0&rdScrollY=0&rdRnd=22817) (Prevalence of hypertension [Htn] in adults >18 years in 2017, US);
http://ask.chis.ucla.edu/AskCHIS/tools/layouts/AskChisTool/home.aspx#/geography (Prevalence of Htn by county, California, 2018)
<sup>5</sup>https://data.oecd.org/health/hospital-beds.htm (Number of beds/1000 population, 2019, OECD); AHA Annual Survey, 2019 by Health Forum, LLC, an affiliate of the American Hospital Association. http://www.ahaonlinestore.com. (Number of beds/1000 population, 2019, US); https://belonging.berkeley.edu/hospital-beds-1000-people-and-risk-populations-county (Number of beds/1000 population, California counties, 2020)

Each regions’ CFR was compared to co-factors that could potentially affect CFRs—date of initiation of the social-distancing measures, median age of the region’s population, prevalence of co-morbidities (diabetes mellitus and hypertension), the number of hospital beds/1000 population (recent data on the number of ICU beds per population were not uniformly available for all regions included in this study), and the number of RT-PCR tests performed/1 million population (Table 1). In Western Europe, the social-distancing measures were initiated on February 21 in Italy and on March 23 in the United Kingdom. In the US, this implementation varied from March 19 to March 24, while in California, San Francisco county was the first to begin the lockdown on March 17, followed by rest of the state on March 19. The timing of the start of these measures showed no correlation with cumulative CFRs. The correlation coefficients comparing cumulative CFRs to median age, diabetes prevalence, hypertension prevalence, number of hospital beds per 1000 population, and number of RT-PCR tests done/1 million population were 0.42, −0.7, −0.47, −0.2, and −0.16, respectively.

We examined 34,412 SARS-CoV2 complete genome sequences deposited from the high-incidence countries and regions in Western Europe, the United States, and California as of June 4, 2020 (Table 2). All of the sequences were compared to that of one of the first SARS-CoV2 strains isolated in Wuhan, China (Wuhan HU-1; MN908947) (1). The number of sequences submitted from each region varied (Table 1, <u>Supplementary Table 1</u>). The United Kingdom submitted the greatest number of sequences (Table 2).

**Table 2.** Cumulative frequencies of the SARS-CoV2 subclades compared to case fatality rates (CFR) in Western Europe, the US, and California counties based on sequences in the GISAID database as of June 4, 2020.

| Country/Region | CFR | Number of sequences analyzed | A23403G (%) | A23403G-C14408T (%) | A23403G-C14408T-G25563T (%) | OR (95% Confidence Interval) <sup>1</sup> |
| --- | --- | --- | --- | --- | --- | --- |
| Germany | **4.7** | 212 | 15 (7.1) | 112 (52.8) | 46 (21.7) | -- |
| Spain | 10 | 967 | 9 (0.9) | 479 (49.5) | 27 (2.8) | 0.88 (0.64-1.19)<sup>2</sup> 0.1(0.06-0.18) <sup>3</sup> |
| UK | 14 | 15,413 | 15 (0.1) | 10,768 (69.9) | 982 (6.4) | 2.1 (1.6-2.7)<sup>2</sup> 0.25 (0.17-0.35)<sup>3</sup> |
| Italy | 14 | 109 | 0 (0) | 99 (90.8) | 2 (1.9) | 8.8 (4.3-20)<sup>2</sup> 0.07(0.01-0.27)<sup>3</sup> |
| France | 15 | 388 | 2 (0.5) | 154 (39.7) | 196 (50.5) | 0.59 (0.41-0.84)<sup>2</sup> 3.8 (2.5-5.5)<sup>3</sup> |
| United States | 5.6 | 6,837 | 4 (0.06) | 675 (9.9) | 4108 (60.1) | -- |
| California | **3.4** | 521 | 0 (0) | 54 (10.4) | 162 (31.1) | -- |
| New York | 7.7 | 1,342 | 0 (0) | 94 (7.0) | 1154 (86.9) | 0.65 (0.45-0.94)<sup>2</sup> 14 (11-17)<sup>3</sup> |
| California county | -- |  |  |  |  |  |
| San Francisco | **1.6** | 154 | 0 (0) | 22 (14.3) | 64 (41.6) | -- |
| Santa Clara | 5.1 | 220 | 0 (0) | 8 (3.6) | 20 (9.1) | 0.23 (0.085-0.55)<sup>2</sup> 0.15 (0.08-0.25)<sup>3</sup> |
| San Diego | 4 | 90 | 0 (0) | 17 (18.9) | 60 (66.7) | 1.4 (0.65-3.0)<sup>2</sup> 2.8(1.6-5.0)<sup>3</sup> |
| China | 5.6 | 742 | 15 (2.0) | 19 (2.6) | 8 (1.1) | -- |

<sup>1</sup>calculated in reference to a country or sub-region with the lowest CFR in the region.
<sup>2</sup>A23403G-C14408T cumulative strain frequency in high vs lowest CFR region.
<sup>3</sup>A23403G-C14408T-G25563T cumulative strain frequency in high vs lowest CFR region.

In Western European countries, the United States, and in California, two of three subclades descending from the Wuhan HU-1 strain in China dominated. One variant had a nonsynonymous mutation in the spike protein gene (A23403G). This subclade contained two additional mutations--one in the leader sequence (C241T) and one synonymous mutation in the gene encoding nonstructural protein nsp3 (C3037T). Descending from this subclade was a variant that gained a missense mutation in a segment of Orf1ab encoding the RNA-dependent RNA polymerase (C14408T). The third subclade contained an additional missense nucleotide substitution in Orf3a (G25563T). These missense mutations led to non-conservative amino acid substitutions in the spike protein (D614G), RNA-dependent RNA polymerase (P323L), and Orf3a protein (Q57H). The remainder of this paper will thus focus on these subclades carrying these missense mutations--A23403G, A23403G-C14408T, and A23403G-C14408T-G25563T.

In Western Europe, the proportion of the A23403G-C14408T subclade varied from 39.7% in France to 90.8% in Italy (Table 2). The cumulative proportions of A23403G-C14408T-G25563T mutant in high CFR Western European countries varied from 1.9% in Italy to 50.5% in France. Italy became the first epicenter of the COVID-19 epidemic in Europe. Of 109 sequences deposited from Italy, 99 (90.8%) carried the A23403G-C14408T mutations, while only 2 (1.9%) carried the A23403G-C14408T-G25563T mutation (Table 2). The A23403G-C14408T subclade was associated with high CFR when the United Kingdom (OR=2.1; 95% CI: 1.6-2.7) and Italy (OR=8.8; 95% CI: 4.3-20) were compared to Germany, but not when Spain and France were compared to Germany (Table 2). The A23403G-C14408T-G25563T subclade was associated with high CFR when France was compared to Germany (OR=3.8; 95% CI: 2.5-5.5), but not when the other Western European countries were compared to Germany (<u>Supplementary Table 2A, 2B, 2C</u>).

In the US, most of the sequence submissions were from New York and California (Table 2). The cumulative frequency of the A23403G-C14408T mutant in New York (high CFR state) was 7%, while in California, it was 10.4% (OR= 0.65; 95% CI: 0.45-0.94). The cumulative frequency of the A23403G-C14408T-G25563T mutant in New York was 86.9% compared to 31.1% in California (OR=14; 95% CI: 11-17). Within California, the proportion of A23403G-C14408T mutant in San Diego vs San Francisco was 18.9% vs 14.3%, respectively (OR= 1.4; 95% CI: 0.65-3.0), while the frequencies of the A23403G-C14408T-G25563T variant were 66.7% vs 41.6%, respectively (OR=2.8; 95% CI: 1.6-5.0). On the other hand, compared to San Francisco, the proportion of A23403G-C14408T subclade in Santa Clara county with a CFR of 5.1% was 3.6% (OR=0.23; 95% CI: 0.085-0.55), and the proportion of A23403G-C14408T-G25563T subclade was 9.1%% (OR=0.15; 95% CI: 0.08-0.25) (Tables 2A-C, Supplementary material).

To determine where these subclades originated, we examined the sequences deposited from China. Of 742 sequences from China deposited mostly from January through March, we found 15 (2%) A23403G, 19 (2.6%) A23403G-C14408T and 8 (1.1%) A23403G-C14408T-G25563T strains (Table 2). Thus, the latter two dominant nucleotide variants in Europe and the US most likely originated in China and greatly expanded in these regions.

The cumulative CFRs and the cumulative frequencies of the dominant subclades we compared above do not reflect temporal changes of these two variables over the course of each epidemic in a region. Thus, we compared the daily number and 7-day moving averages of the frequencies of these mutants to the daily number of cases and deaths across the course of each epidemic. This analysis had to be limited to samples from countries and regions that submitted a sufficient number of sequences from the early phase to shortly after the peak period of the epidemic.

In the United Kingdom, among 15,413 full genome sequences analyzed, the 7-day moving average of the frequencies of the A23403G-C14408T subclade increased from less than 30% at the beginning of the epidemic in late February to over 90% just past the peak of the epidemic (Fig. 1). bThe 7-day moving average of the A23403G-C14408T-G25563T variant remained below 10% throughout most of the epidemic period. In Germany, from late February to mid-March, the 7-day moving average of the proportions of A23403G-C14408T mutant increased quickly to above 70%, dipped briefly to just above 40%, and then increased again to over 80% just past the peak of its epidemic in late April (Fig. 1). The A23403G-C14408T-G25563T subclade appeared to compete with the other subclade, initially peaking at nearly 50%, then falling to less than 10% by the post-peak period. The two subclades’ 7-day moving averages moved in opposite directions, where the double-mutant seemed to outcompete the triple mutant (Fig. 1). The same phenomenon is observed in France (Fig. 1). In Germany, the number of deaths peaked in mid-April, about a month after the peak 7-day average proportion of the A23403G-C14408T-G25563T strain. In both UK and Spain, the A23403G-C14408T-G25563T subclade’s 7-day moving average remained at less than 10% throughout their epidemics.

**Figure 1.**
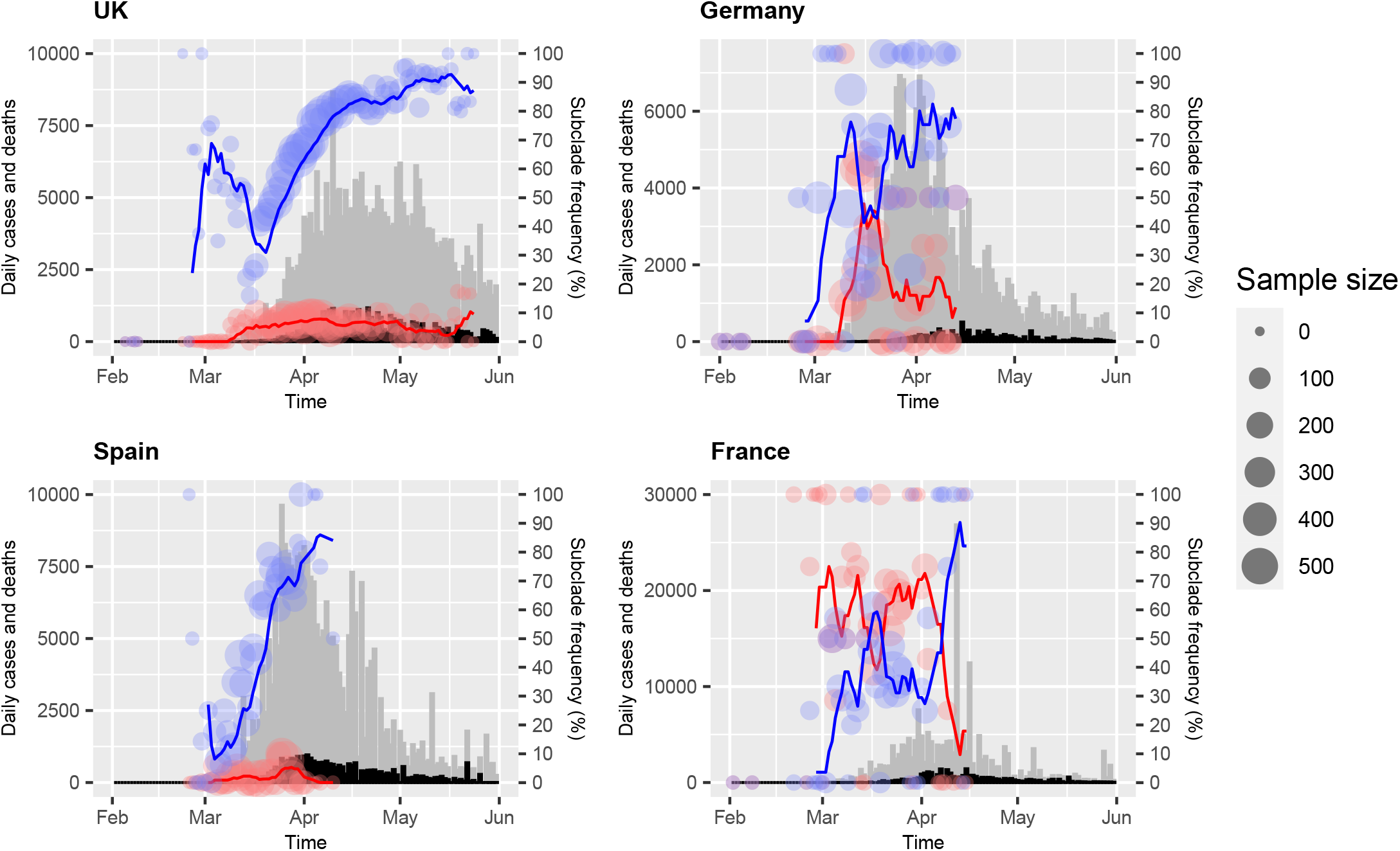
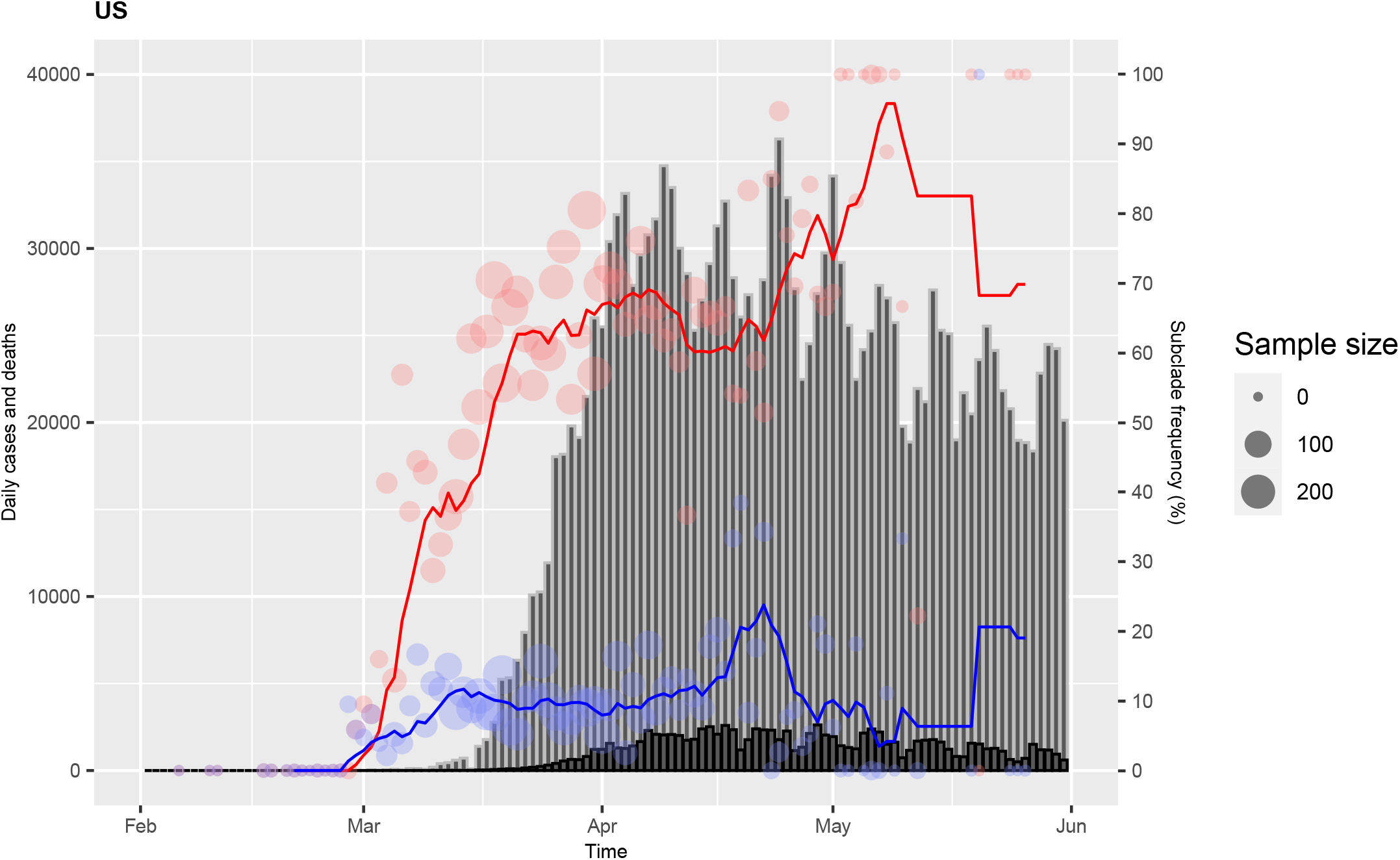
Daily numbers (filled circles) and seven-day moving average of the frequencies of A23403G-C14408T subclade (blue line) and C14408T-A23403G-G25563T subclade compared to cases (gray bar) and deaths (black bar) in western European countries that submitted > 200 SARS-CoV2 genome sequences. In Germany that had a cumulative case fatality rate of 4.7% at the end of first week of June, both subclades peaked at the beginning of the epidemic, which preceded the peak of deaths by 3-4 weeks. Deaths appear to have decreased after the A23403G-C14408T-G25563T subclade decreased.

Interestingly, at the beginning (February) of the epidemic in Germany, more than 90% of the strains carried the A23403G mutation but not the C14408T or G25563T mutations. Strains with C14408T or G25563T mutation were not observed during this early period. The A23403G single nucleotide variant was completely replaced by mid-March by the other two subclades. In all the other regions in Western Europe and the US, this single nucleotide variant represented less than 1% of the sequences deposited in the GISAID database (Table 2).

In the United States, unlike Europe, it was the A23403G-C14408T-G25563T mutant that became the dominant strain (Fig. 2). In New York, the state that had the highest cumulative CFR, the epidemic (cases and deaths) peaked in early April. Of 1342 complete sequences submitted from the state, the moving 7-day average of the proportions of the A23403G-C14408T mutant fluctuated between 0 and 20% during most of the epidemic course (Fig. 2). During the same period, the 7-day moving average of the A23403G-C14408T-G25563T subclade fluctuated around 90% (Fig. 2). Interestingly, in California that had a relatively low cumulative CFR, the epidemic started with the triple mutant clade, but by mid-May, the A23403G-C14408T mutant took over.

**Figure 2.**
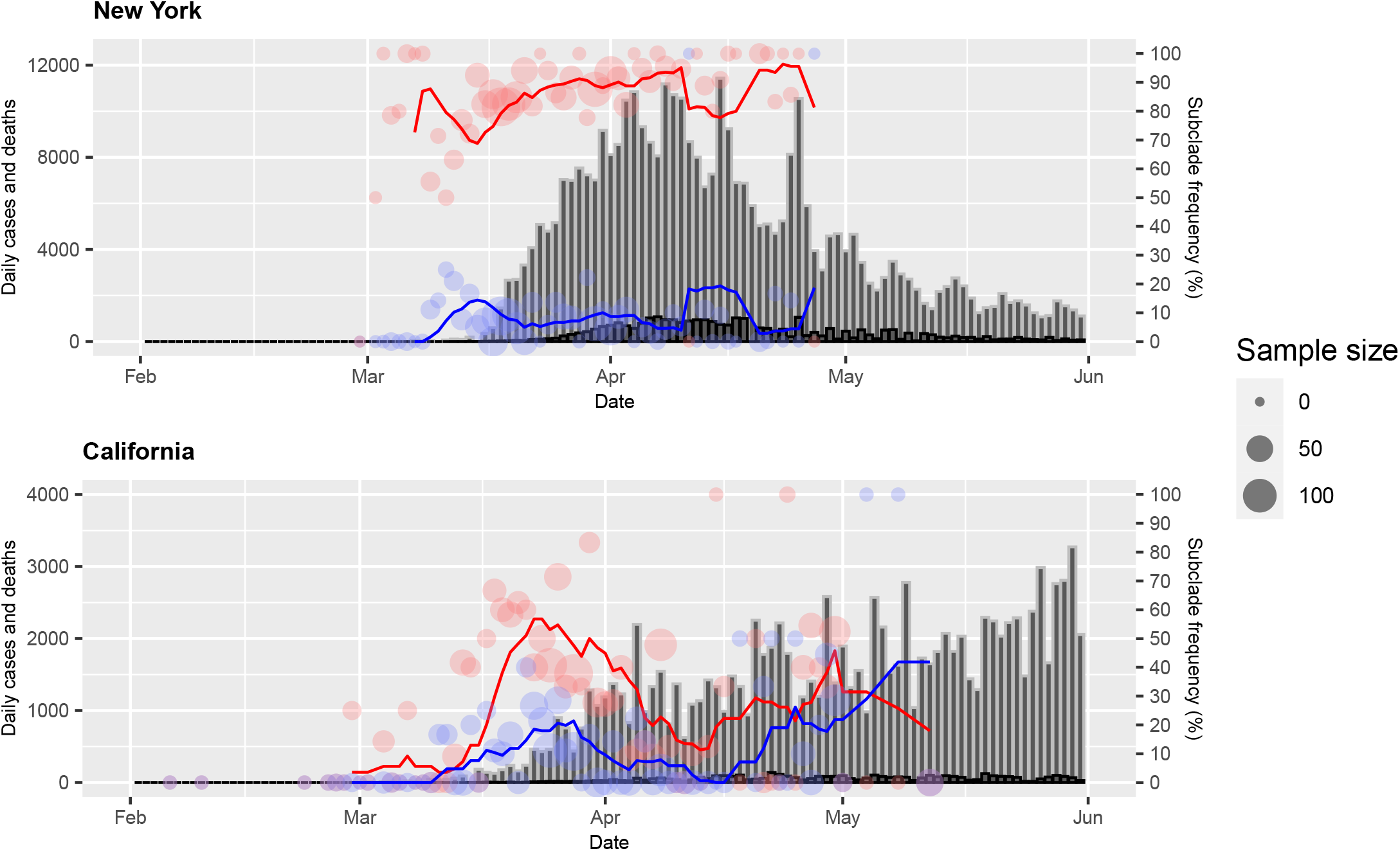
Daily numbers (filled circles) and seven-day moving average of the frequencies of A23403G-C14408T subclade (blue line) and A23403G-C14408T-G25563T subclade (red line) compared to cases (gray bar) and deaths (black bar) in the United States (2A), New York, and California (2B) that submitted > 500 SARS-CoV2 genome sequences. Unlike Europe, in the United States, the A23403G-C14408T-G25563T subclade appears to be dominant.

Within California, Santa Clara county had low frequencies of both mutants, despite having a high CFR relative to San Francisco county. However, Fig. 3 shows that, as observed in Germany, Santa Clara county actually had high frequencies of both of these mutants during the early phase of the epidemic (March) and both mutants decreased rapidly to less than 5% by early April. Nearly 45% of the deaths in this county occurred in long-term care facilities (LTCFs). The peak number of deaths that were not associated with LTCFs occurred in late March (https://www.sccgov.org/sites/covid19/Pages/dashboard-cases.aspx), shortly after the peak 7-day mean average period of the two mutants (Fig. 3).

**Figure 3.**
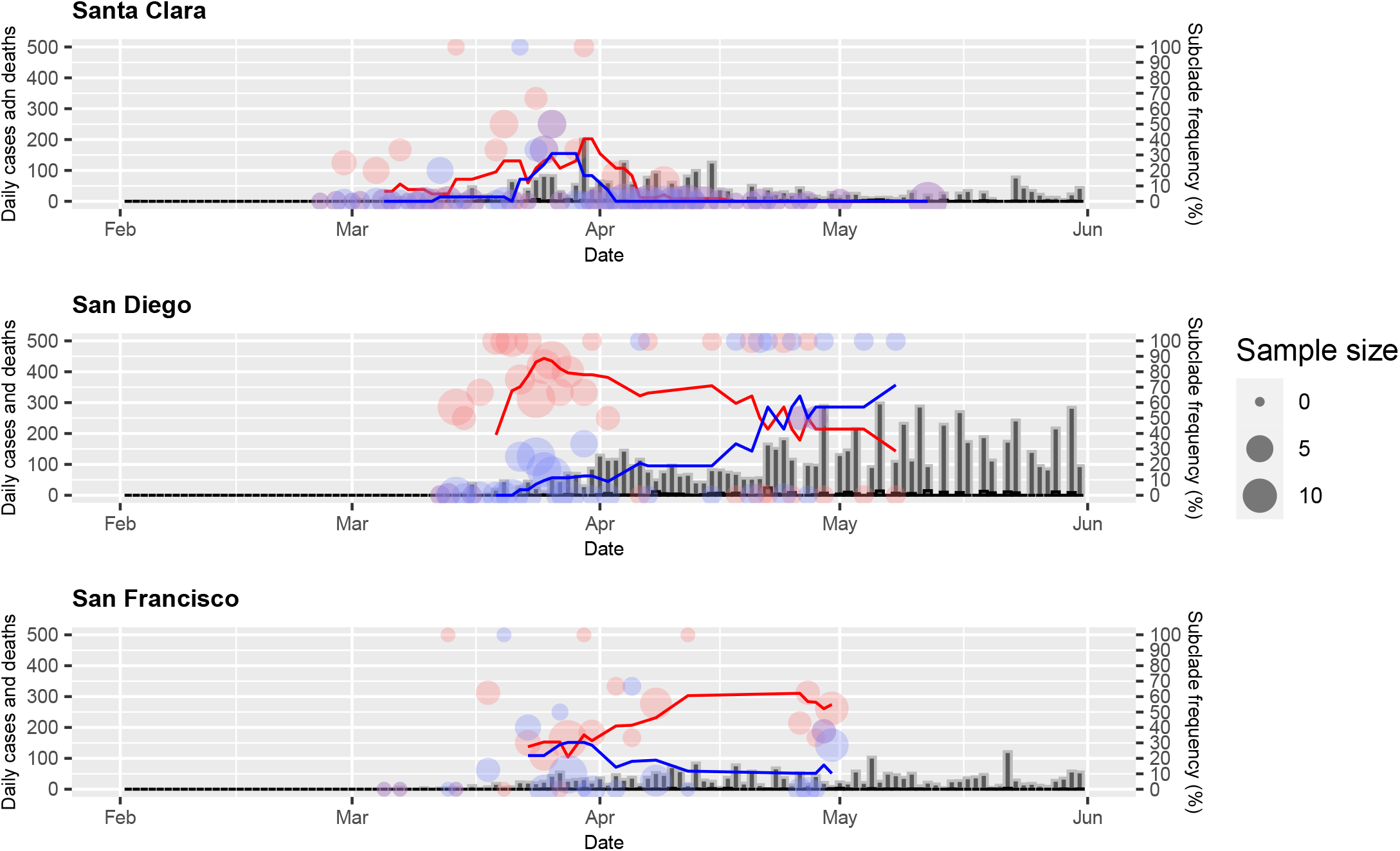
Daily numbers (filled circles) and seven-day moving average of the frequencies of A23403G-C14408T subclade (blue line) and A23403G-C14408T-G25563T subclade (red line) compared to cases (gray bar) and deaths (black bar) in three counties in California (San Francisco, San Diego, and Santa Clara) that submitted >90 SARS-CoV2 genome sequences. Santa Clara county had the highest cumulative CFR while the cumulative frequencies of the subclades were lowest of the three counties. Nearly half of the deaths in Santa Clara county involved residents of long-term care facilities (LTCFs). Note that the two subclades dominated at the beginning of the epidemic, which preceded the peak period of deaths in people who were not residents of LTCFs.

## Discussion

This genomic analysis of 34,412 SARS-CoV2 sequences deposited in the GISAID database found two dominant subclades of the virus co-circulating in Europe and the United States derived from the Wuhan HU-1 strain isolated in China in December, 2019 (1). These subclades descended from another subclade with a missense mutation in the spike protein gene A23403G (D614G). The A23403G, A23403G-C14408T, and A23403G-C14408T-G25563T variants accounted for only 2%, 2.6%, and 1.1%, respectively of 742 sequences deposited from China as of June 4 (Table 2). The epidemic in Germany initially involved the A23403G variant, which was replaced during the epidemic by the A23403G-C14408T and A23403G-C14408T-G25563T variants. In Italy, which became the first epicenter of the epidemic in Europe, over 90% of the cases were due to the double mutant. However, this observation could have resulted from sampling bias. The small number of samples submitted from the country (109) may not be representative of Italy’s cases. (Table 1, Supplementary Material).

The dominant subclades were significantly associated with some of the high cumulative CFR regions in Western Europe, the United States, and in California, even after controlling for differences in the initiation of social distancing measures, population age structure, prevalence of diabetes mellitus or hypertension, number of hospital beds/1000 population, and number of RT-PCR tests/1 million population done. Among five Western European countries reporting >150,000 cases at the end of May 2020, Germany had the lowest cumulative CFR (4.7%). Yet, the lockdown in Germany was initiated after Italy, Spain, and France. Germany’s median age (47.8 years) is the highest, and it has the highest prevalence of diabetes and second highest prevalence of hypertension among these countries. It does boast the highest number of hospital beds per 1000 population, but this factor did not correlate with low CFR in the other four European countries, the US or California counties (Table 1).

The cumulative frequencies of A23403G-C14408T and A23403G-C14408T-G25563T variants were not each consistently associated with the high cumulative CFR regions. The A23403G-C14408T mutant was significantly associated with high CFR when the United Kingdom and Italy were compared to Germany. On the other hand, the A23403G-C14408T-G25563T mutant was associated with high CFR only when France was compared to Germany. In the US, it was the triple nucleotide mutant that was associated with high CFRs. In early June, the CFR in New York was 7.7%--more than double that of California at 3.4%, and the A23403G-C14408T-G25563T mutant was significantly more prevalent in New York (Table 2). Within California, this mutant was significantly associated with high CFR when San Diego county was compared to San Francisco county. However, this association was not observed with Santa Clara county, which had a high CFR at 5.1% but low frequencies of both subclades at less than 10%.

When the frequencies of the variants were examined by their 7-day moving averages, a more consistent pattern was observed. In Western Europe, interestingly, it was the A23403G-C14408T mutant that dominated in most of the countries. However, the peak periods of deaths appeared to temporally correspond to the period of increased co-circulation of the A23403G-C14408T-G25563T mutant (Fig. 1). In Germany, when we examined the frequency of the mutants by intervals of 7-day moving averages over the course of the epidemic, the A23403G-C14408T-G25563T mutant peaked at close to 50% during the early phase of the epidemic, which was about 3-4 weeks before the peak number of deaths (Fig. 2). Since the virus isolation is most likely made at the time of initial diagnosis of COVID-19, if the co-circulation of the two mutants is associated with increased number of deaths, it would be expected that the peak 7-day mean average of the co-circulating mutants will precede the peak number of deaths during the epidemic.

In the United Kingdom, the cumulative proportion of the A23403G-C14408T-G25563T mutant was low (6.4%) but the UK’s CFR was one of the highest in Europe at 14%. The UK reported more than 255,000 cases and 40,000 deaths at the end of first week of June. Thus, even if only 6.4% of the people were infected with this strain, more than 16,000 people would have been infected, suggesting that both of the subclades were co-circulating at high numbers. Thus, in regions where the case numbers are high, the numbers of these circulating dominant mutants would also be high, even if the relative proportion of one may be low. Part of the high mortality rate of COVID-19 observed in the UK, therefore, could still be related to the high level of co-circulation of these dominant strains (Fig. 2, Table 2).

In California, Santa Clara county reported a high cumulative CFR (5.1%) relative to San Francisco at 1.6%. The two mutants together accounted for over 90% of the strains circulating in Santa Clara in late March but both fell rapidly to less than 5% by early April (Fig. 3). It should be noted that this county reported less than 3000 cases and 150 deaths at the end of first week in June. Nearly half of all deaths in the county through May occurred in long-term care facilities (LTCF). Thus, the CFR was greatly affected by these outbreaks at LTCFs. Interestingly, the peak 7-day average of the two mutants occurred 2-3 weeks prior to the peak number of non-LTCF deaths, again suggesting that the co-circulation of the two subclades may contribute to increased mortality.

In San Francisco with a CFR of only 1.6%, the triple mutant dominated. However, it fluctuated at levels between 20 to 60%--less than what was observed in high CFR city like New York City, which had a CFR of 10%. The small number of samples available from San Francisco (154) could have also contributed to bias in the representativeness of the samples.

It is well recognized that most of the severe complications or deaths due to COVID-19 occur in those who are over 65 years of age or those with co-morbidity (8, 9). But the mechanisms that lead to the respiratory failure accompanied by hyperinflammatory state in these patients are unknown. The co-circulation of dominant SARS-CoV2 subclades raises a disturbing possibility that people living in places with high prevalence of co-circulating strains may get serially infected with each variant. The second infection may trigger an immune response not unlike the antibody-dependent enhancement (ADE) observed in diseases such as dengue. In settings where a population gets serially exposed to different strains of the dengue virus, the exposed individuals may mount varying levels of heterotypic antibody response directed at the different viral strain that do not neutralize but enhance binding of the strain to enter host cells (10). Infected with SARS-CoV2, those with underlying medical conditions are likely to mount lower levels of neutralizing antibody response to the first infection, putting them at risk for ADE-like response during the second infection.

Indeed, evidence for spike protein antibody-mediated viral entry into cultured cells has been demonstrated for the Middle East respiratory syndrome (MERS) coronavirus (11). A potential for ADE-like host response from natural serial infections with the dominant SARS-CoV2 mutants may need to be considered for COVID-19 vaccine development.

The non-conservative amino acid changes that occurred in the SARS-CoV2 clades involve proteins that have each been suggested to affect pathogenicity and transmissibility of the virus. The SARS-CoV2 Orf1ab encodes a number of non-structural proteins, including RNA dependent RNA polymerase (RdRp) or nsp12 (12). The nonsynonymous nucleotide substitution C14408T in Orf1ab leads to P323L amino acid substitution in RdRp (12). The G25563T mutation in Orf3a leads to a non-conservative amino acid change Q57H. In a study of 2,782 SARS-CoV-2 sequences downloaded from the GISAID database in early April, 51 different nonsynonymous amino acid substitutions in the Orf3a protein were found (13). The isolates defined by these mutations were separated into distinct subpopulations according to six functional domains. These functional domains were suggested to be associated with infectivity, ion channel formation, and virus release (13). In SARS-CoV1, the Orf3a protein was reported to be a virulence factor that binds to TRAF3 to activate NF-κB and the NLRP3 inflammasome (14). If the SARS-CoV2 Orf3a protein can exhibit a similar host response, it is biologically plausible that the Q57H mutation in Orf3a could contribute to the severe clinical manifestations observed in COVID-19.

The A23403G mutation in the spike protein gene leads to a non-conservative amino acid change D614G. The spike proteins of SARS-CoV1 and CoV2 share a receptor-binding domain (RBD) that facilitates virus entry into mammalian cells via the angiotensin converting enzyme-2 (ACE2) receptor (15, 16). ACE2 is also the receptor for a group 1 coronavirus NL63, which causes common cold (17). Thus, cell entry, while required, does not appear to be sufficient for SARS-CoV2 to cause the severe hyperinflammatory clinical manifestations. Nevertheless, as recently reported by Korber et al, the mutation D614G in the spike protein could have provided a selective advantage that enabled these two subclades to become the dominant strains in Europe and the United States (3). Indeed, another report showed that the SARS-CoV2 G614 spike protein is more stable than the D614 protein and that a pseudovirus (based on Maloney murine leukemia virus) expressing G614 was able to infect human ACE2-expressing HEK293T cells more efficiently than that expressing the D614 protein (18). Korber et al also showed that patients infected with the D614G mutant had a higher viral load and that infectious titers of vesicular stomatitis virus (VSV) and lentiviral particles expressing the spike protein with the G614 mutation was higher than those expressing the D614 mutation (3). In a sub-analysis of COVID-19 patients in the United Kingdom, whose viral sequences were available from the GISAID database, they found that infection with the D614G mutant was not associated with increased hospitalization, although the mutant was slightly but not significantly enriched among the ICU patients (3).

The impact of the mutation C14408T in RdRp on SARS-CoV2 viral function or virulence is unknown, but strains carrying this mutation were shown by others to have a higher number of co-mutations than strains without this mutation (12). The mutation in RdRp may have affected the proof-reading function of this enzyme. The dominant clade of this virus with this Orf1a mutation may thus continue to accumulate mutations as it spreads into new regions. In SARS-CoV1, Orf1a has also been reported to facilitate viral adaptation to new hosts (20). If this function exists for SARS-CoV2, this could also explain its highly successful transmissibility in human populations.

The limitation of the findings of our study is that they are based on sequences deposited in the GISAID database, most of which do not have patient clinical data. Hence, the associations we found between high CFRs and co-circulation of the dominant SARS-CoV2 lineages are an ecologic observation. In addition, the number of sequences submitted from each country and region vary greatly and may not necessarily be representative of the strains circulating in these places. Nevertheless, it is striking that the two subclades of the virus were consistently observed to dominate in Europe and the United States. A similar observation was reported by others based on analyses of the sequences deposited in the GISAID database from earlier periods (3, 12, 13).

## Conclusions

Over the course of 6 months of the COVID-19 pandemic, two subclades descending from the Wuhan HU-1 strain became dominant in Europe and the United States. The co-circulation of the two variants in high numbers correlated temporally with periods of high mortality in Western Europe, the United States, and California counties. We speculate that the severe COVID-19 clinical manifestations and deaths could result from aberrant host immune responses to sequential infections in the same person with these subclades. These observations will need to be confirmed with studies linking patient clinical outcome data with the patient’s viral sequences. As the COVID-19 pandemic expands into the southern hemisphere, we may be able to track these subclades of SARS-CoV2 to determine how they affect the shape and magnitude of the epidemic and mortality in each region. Furthermore, in addition to the spike protein, the other viral proteins containing the observed mutations should be considered for detailed investigation for their role in pathogenicity and transmissibility, as well as potential targets for a new vaccine or anti-viral drugs.

## Methods

Cumulative and daily number of COVID-19 cases and deaths in Western Europe and the United States:

Cumulative COVID-19 cases and deaths from Western Europe (United Kingdom, Germany, Spain, France, and Italy), the US (California, Illinois, New York, New Jersey, Michigan), and California counties (San Francisco, Santa Clara, and San Diego) reported in the Worldometer database (https://www.worldometers.info/coronavirus/) as of June 6, 2020 were used to calculate cumulative case fatality rates (CFR). Western Europe countries reporting >150,000 cases and the US states reporting >100,000 cases were selected. The selected California counties included those that submitted > 90 SARS-CoV2 genome sequences into the Global Initiative for Sharing All Influenza Data (GISAID) database. Information on co-factors potentially affecting CFRs in these regions (lockdown initiation date, median age of cases, prevalence of diabetes mellitus, prevalence of hypertension, number of hospital beds per population, and number of RT-PCR tests per population) were obtained from sources indicated under Table 1. Correlation coefficients of these variables with CFR were calculated with R software, version 3.6.3.

Daily COVID-19 case and death numbers from Western European countries (United Kingdom, Germany, France, Spain, and Italy) and daily COVID-19 case and death numbers from the US states (New York and California) and California counties (San Francisco, Santa Clara, and San Diego) were obtained from the repository (https://github.com/CSSEGISandData/COVID-19) hosted by the Center for Systems Science and Engineering (CSSE) at Johns Hopkins University (19). Data were accessed on June 4, 2020.

### SARS-CoV2 genome sequence analysis

We initially acquired full sequence alignments of 37,647 submissions to EpiCoV from the Global Initiative for Sharing All Influenza Data (GISAID) database (“msa_0604.fasta”, downloaded on June 4, 2020). Duplicate and low-quality sequences (>5% NNNNs) were removed and alignment of 34,417 sequences was created based on the sequence multiple alignment program MAFFT (20) by GISAID. We analyzed only complete sequences (length >29000 bp). We trimmed the full-length alignment with trimAL3 v1.4.rev15 (21), which has the automated option-automated1 to remove any spurious parts of the alignment, thus reducing noise introduced into the single nucleotide variant analysis. The aligned genomes were re-positioned according to the reference SARS-CoV-2 genome (GenBank access number: NC_045512.2; Wuhan HU-1; MN908947).

We extracted single nucleotide polymorphism (SNP) information with the program snp-sites with default option (22). SNPs with prevalence less than 0.3% among all sequences were excluded from the analysis. All variant information was merged to metadata information based on gisaid_epi_isl number, downloaded on June 4, 2020 from GISAID (“metadata_2020-06-04_18-12.csv”).

### Statistical analysis

The statistical analysis was performed with R software, version 3.6.3. We compared the frequencies of the dominant viral subclade sequences in two categories based on CFR of the region from which they were submitted—high CFR vs low CFR region. In Western Europe, Germany was set as the low mortality region (CFR = 4.7%) and all other countries were classified as high mortality region (CFR > 10%). In the US, low mortality states included California, Illinois, Texas, Florida, and Georgia (case-fatality rates, CFR < 5%), and high mortality states included New York, New Jersey, Michigan, Connecticut, and Louisiana (CFR). In California, the frequency of the sequences of the isolates from San Francisco (CFR of 1.6%) was compared to that of San Diego, which had a cumulative CFR of 4% as of June 6. Frequencies of dominant subclades were compared between high CFR and low CFR regions by Fisher’s exact test. All p-values were calculated from 2-sided tests based on 8.8e-06 as the significance level based on Bonferroni correction for multiple comparisons.

### Subclade analysis

All analysis was conducted with R package dplyr (23) and graphically plotted with ggplot2 (24). Odds ratios and their 95% confidence intervals of cumulative strain frequencies of the dominant strains were calculated in reference to a country or sub-region with the lowest CFR in the region, using Fisher’s exact test. Since CFRs and strain frequencies fluctuate by time, we also examined daily frequencies and 7-day moving averages of the frequencies of the dominant subclades by plotting them against daily cases and deaths. Peak frequencies of the subclades were compared to peak numbers of death during each epidemic.

## Data Availability

1. The number of SARS-CoV2 genome sequences from each country in the GISAID database that were included in the analyses are provided in the Supplementary material Table 1.
2. Calculations of Odds Ratio and 95% CI to assess association of the dominant subclades with high case-fatality rates (CFRs) are provided in Supplementary material Tables 2A-C.
3. URLs of the sources of data regarding co-factors that may affect CFRs are shown under Table in the manucript. 

https://www.worldometers.info/coronavirus/

https://www.gisaid.org/

https://www.sccgov.org/sites/covid19/Pages/dashboard-cases.aspx

https://www.nbcnews.com/health/health-news/here-are-stay-home-orders-across-country-n1168736

https://www.cia.gov/library/publications/the-world-factbook/fields/343.html

https://worldpopulationreview.com/

https://www.diabetesatlas.org/upload/resources/2019/IDF_Atlas_9th_Edition_2019.pdf

https://gis.cdc.gov/grasp/diabetes/DiabetesAtlas.html#

https://www.who.int/nmh/countries/

https://gis.cdc.gov/grasp/diabetes/DiabetesAtlas.html#

## Author Contributions

LR conceived of the idea to compare regional differences in CFR with viral variants, obtained co-factor data, and calculated regression coefficients. YH accessed the GISAID database, performed the genome sequence analyses and graphically presented the data. LR and YH interpreted the findings and both prepared the manuscript.

